# The effect of BCG vaccination on adult mortality during the COVID-19 pandemic: a meta-analysis of randomised controlled trials

**DOI:** 10.1101/2025.11.19.25340570

**Authors:** Frederik Schaltz-Buchholzer, Nigel Curtis, Andreas H. Diacon, Evangelos J. Giamarellos-Bourboulis, Antigone Kotsaki, Anne Marie Rosendahl Madsen, Nicole L. Messina, Lidia Nhamússua, Sebastian Nielsen, Laure F. Pittet, Isaquel da Silva, Annie Sparrow, Caryn Upton, C.H. van Werkhoeven, Mihai G. Netea, Christine S. Benn, Peter Aaby

## Abstract

**Objectives:** Bacillus Calmette-Guérin (BCG), similar to some other live vaccines, may offer partial protection against unrelated infections. During the COVID-19 pandemic, several trials tested whether BCG provides protection against COVID-19.

**Design:** Meta-analysis of randomised placebo-controlled trials which provided mortality data.

**Setting:** High-income settings as well as low- and middle-income settings (LMICs).

**Participants:** Trial populations were healthcare workers (HCW) or individuals of older age, followed for 6-12 months.

**Interventions:** BCG vaccine versus placebo.

**Main outcome measure:** Overall mortality and COVID-19 mortality.

**Results:** Nine of the 15 RCTs reported at least one death. Trial populations were healthcare workers (4 RCTs) or individuals of older age (5 RCTs), in Europe (5 RCTs) or LMICs (4 RCTs) with a total of 8,169 participants randomly allocated to BCG (18 deaths) and 8,176 to placebo (37 deaths). In a meta-analysis, receiving BCG vs placebo was associated with a 51% (95% CI: 15 to72%) reduction in all-cause mortality. The all-cause mortality RRR was 78% (−3 to 95%) in HCWs (Europe and LMICs) and 43% (95% CI: −6 to 69%) in the elderly. It was 37% (95 CI: −15 to 66%) in Europe and 90% (95% CI: 22 to 99%) in LMICs. The relative reduction of deaths (6 BCG; 12 placebo) from COVID-19 was 50% (95 CI: −33 to 81).

**Conclusions:** BCG may have reduced all-cause mortality in the context of the COVID-19 pandemic.

## Introduction

The current public health paradigm has a strong focus on identifying the pathogens causing health problems and on developing specific treatments and/or vaccines against these pathogens. However, strategies that broadly strengthen the immune response could also contribute to decreasing the susceptibility to both existing and new diseases and/or reducing the severity of such diseases (1–4).

During the last three decades, many studies, mainly in infants, have suggested that live attenuated vaccines may have beneficial non-specific effects (NSEs), reducing susceptibility or severity of non-targeted infections (1). This has been shown for BCG (5,6), measles vaccine (MV) (7), measles-mumps-rubella vaccine (MMR) (8), oral polio vaccine (OPV) (9), and *Vaccinia* against smallpox (1). For BCG, MV, and OPV, non-specific (off-target) benefits have been documented in randomised controlled trials (RCTs) (5–7,9–10). In these, the NSEs of live vaccines have been significant, reducing all-cause/infectious disease mortality by 30-40% in high-mortality settings (5–7,9–10). These effects may be related to the capacity of live vaccines to reprogram the function of the innate immune system through epigenetic modifications, a process known as “trained immunity” (3).

With benefits of NSEs demonstrated both at the epidemiological and immunological level, live vaccines may be useful as a tool to strengthen the first line of defense when a new “*Disease X*” with pandemic potential emerges (2,11). A non-specific vaccine that mitigates the negative health impact of a new infection could have significant public health and societal impact.

When COVID-19 emerged as a pandemic in 2020, the focus was to develop specific treatments and specific vaccines. In parallel, however, several groups suggested that live attenuated vaccines could be used as stopgap vaccines until specific vaccines became available (2,11). Subsequently, BCG (12–20), OPV (21,22) and MMR (23) were tested in RCTs as an intervention against COVID-19. Most RCTs tested BCG against COVID-19.

The results of these RCTs are now available. The effect of BCG against the incidence of COVID-19 has been inconsistent: while some of these RCTs suggested that BCG may exert moderate beneficial effects, most studies showed no beneficial effects against total number of COVID-19 episodes or increased incidence of infections (19,24–26).

A critically important outcome in a pandemic situation is, however, all-cause mortality. Early in the pandemic, it was hypothesised that BCG might have a beneficial effect on adult survival (2,11), as has been previously documented for child mortality (5,6). Here, we review the results of the RCTs of BCG vs placebo against COVID-19 for their impact on mortality.

## Methods

### RCTs of BCG

In November 2024, we searched PubMed for RCTs of BCG vaccination against COVID-19 as the main outcome (N=70). Since we were only interested in the effects during the SARS-CoV-2 pandemic, we searched for studies between the years 2020-2024 with the search input: *((randomized or randomised) trial or RCT) and (BCG or Bacille Calmette Guerin) and (COVID-19 or SARS-CoV-2).* All studies investigating the effect of BCG on SARS-CoV-2 related outcomes, irrespective of BCG strain, were included.

In the main analysis, we excluded one study that used VPM1002, as this is a genetically modified BCG, which also expresses listeriolysin, and which might therefore have a different effect on trained immunity. However, we included this study in a sensitivity analysis.

Only RCTs which reported deaths by randomisation group in the main publication were included in the main analysis (Table 1); other RCTs without deaths have been listed in Supplementary Table 1. For one RCT, which recruited participants from multiple countries, only the countries for which deaths were reported were included in the main analysis.

**Table 1.** RCTs of BCG Vaccine versus Placebo against COVID-19 with one or more death during follow-up.

| Country;<br>Authors | Study population;<br>Age | BCG strain | Follow-up<br>period | Effect on main<br>outcome | All-cause mortality<br>(Deaths/N) [Death<br>from COVID-19] |  | All-cause<br>mortality risk<br>ratio<br>(BCG/placebo)<br>(95%CI)<br>Fixed effects |
| --- | --- | --- | --- | --- | --- | --- | --- |
|  |  |  |  |  | BCG | Placebo |  |
| RCTs against COVID-19 in Europe |  |  |  |  |  |  |  |
| Greece (12) | Elderly with co-morbidity, at hospital discharge; 50+ years | BCG-Moscow | 6 months | 2 patients in BCG and 10 in placebo group | 0/148 | 3/153<br>[1 COVID-19] | 0 (undefined) |
| The Netherlands (13) | Elderly with co-morbidity; 60+ years | BCG-Denmark | 6 months (182 days after enrolment) | HR of COVID-19 episode 1.12 (0.87-1.44));<br>HR of COVID-19 hospitalisation 0.86 (0.46-1.61) | 13/3058<br>[5 COVID-19] | 18/3054<br>[6 COVID-19] | 0.72 (0.35-1.47) |
| The Netherlands (14) | Healthy elderly; 60+ years | BCG-Denmark | 12 months | HR of respiratory infections 1.26 (0.65-2.44);<br>HR of COVID-19 episode 1.05 (0.71-1.56) | 2/1008<br>[1 COVID-19] | 3/1006 | 0.67 (0.11-3.97) |
| The Netherlands (15) | Healthcare workers; Median age: 42 years | BCG-Denmark | 12 months | RR of absenteeism 0.94 (0.78-1.15);<br>RR of documented COVID-19 0.94 (0.72-1.24) | 1/753 | 0/758 | Infinite (Undefined) |
| Denmark (16) | Healthy elderly; 65+ years | BCG-Denmark | 12 months | HR for acute infection 0.89 (0.78-1.02) | 1/838 | 3/838 | 0.33 (0.03-3.20) |
| Europe |  |  |  |  | 17/5805 | 27/5809 | 0.63 (0.34-1.16) |

| RCTs against COVID-19 in LMIC |  |  |  |  |  |  |  |
| --- | --- | --- | --- | --- | --- | --- | --- |
| **India (17)** | High-risk adults (diabetes, chronic kidney, liver, or lung disease); Mean age: 44 years | BCG-Moscow | 9 months | 62% (28-80%) reduction in likely symptomatic COVID-19 | 0/246 | 1/249<br>**[1 COVID-19]** | 0 (undefined) |
| **South Africa (18)** | Healthcare workers; Median age: 39 years | BCG-Denmark | 12 months | HR of COVID-19: 1.08 (0.82-1.44)<br>HR of hospitalisations 2.00 (0.69-5.9) | 0/500 | 4/500<br>**[2 COVID-19]** | 0 (undefined) |
| **BRACE Brazil (19)** | Healthcare workers with no prior COVID-19; Median age: 42 years | BCG-Denmark | 12 months | All-cause mortality result (including participants in Europe and Australia): 3% (0.2-5.8%) higher risk of COVID-19 episode; Hospitalisations HR 1.56 (0.60-4.02) | 0/1285 | 4/1283<br>**[2 COVID-19]** | 0 (undefined) |
| **Guinea-Bissau, Mozambique (20)** | Healthcare workers; Mean age: 34 years | BCG-Denmark | 6 months | HR of absenteeism 1.29 (0.81-1.94); HR of hospitalisation 0.51 (0.13-2.09) | 1/333 | 1/335 | 1.01 (0.06-16.02) |
| **LMIC** |  |  |  |  | 1/2364 | 10/2367 | 0.10 (0.01-0.78) |
| **RCTs of BCG against COVID in Europe and LMIC** |  |  |  |  | **18/8169** | **37/8176** | **0.49 (0.28-0.85)** |

Notes: All trials provided a single dose of BCG, either as primary vaccination or revaccination. RCTs of BCG conducted in the US, Denmark, Poland, Mexico, Brazil and the Australian and European parts of the BRACE trial (19) had no deaths and have been presented in supplementary table 1.
Abbreviations: BCG, Bacille Calmette-Guérin; CI, Confidence Interval; HR, Hazard Ratio; LMIC, Low-and-Middle Income Country; RCT, Randomized Controlled Trial.

We contacted the authors to verify the deaths reported in the paper, the causes of death, particularly whether anyone had died of COVID-19, and whether the participants who died had received COVID-19 vaccine before death. The authors of all RCTs responded, except for one group (17). Most authors were not able to provide complete and certain information on whether the participants who died had received COVID-19 vaccine during follow-up. COVID-19 vaccination data was therefore not analysed.

The data used for this paper are available in the published literature, except where it is indicated that information was provided directly by authors.

### Statistical methods

For most RCTs, we only had access to the numbers of deaths and participants by randomisation group. We therefore analysed the relative mortality risk (RR) of BCG vs. placebo. There was insufficient information to analyse data by sex. The absolute risks for BCG and placebo groups are presented with the number of events as X% (Table 1). Risk ratios with 95% confidence intervals (CI) were calculated using the *metan* command in Stata BE version 18 providing Mantel-Haenszel (MH) estimates. The effect estimate was reported as a “relative risk reduction”, calculated as RRR = (1 – RR) * 100%. The *metan* command was also used to calculate combined MH fixed effects estimates, as recommended over random effects estimates in the case of rare events (31).

## Results

### Study populations

In the PubMed search, we identified nine RCTs (12–20) that investigated the impact of BCG vaccination on COVID-19, and which had deaths recorded during the follow-up period of the trial (Supplementary Figure 1). A review of seven meta-analyses, which covered many different COVID-19 related outcomes, identified no further relevant RCT (4,24,26,32–35). Of the nine RCTs, five were from Europe (Greece (n=1), The Netherlands (n=3), Denmark (n=1)), three were from low-and-middle income countries (LMICs) (South Africa (n=1), India (n=1), and Guinea-Bissau/Mozambique (n=1), and one RCT (BRACE) enrolled participants from Australia, Europe, and Brazil. As there were no deaths in the BRACE trial in participants in Europe and Australia, only the Brazil part of the trial has been included in Table 1. The study populations were either healthcare workers (HCW) or individuals of older age (see Table 1), two of the groups considered most at risk at the onset of the pandemic.

Only the RCTs with at least one death in either the BCG or the placebo group contribute to the estimation of the impact on mortality (36). The RCTs of BCG where there was no death in any group have been listed in Supplementary Table 1 (19,37–42).

### All-cause mortality

The RCTs included 16,345 adults (BCG 8,169; placebo 8,176) among whom there were 55 deaths: 18 in BCG-vaccinated participants; 37 in the placebo-vaccinated participants (Table 1). The meta-analysis effect estimate for the RRR of BCG compared with placebo for all-cause mortality was 51% (15 to 72%). The effect was less marked in Europe, where the age of participants was higher (most RCTs had enrolled elderly, and more participants died of chronic diseases or cancers). There was a significant beneficial effect of BCG vs placebo in the LMICs, the all-cause mortality RRR being 90% (22 to 99%) (1 versus 10 deaths). The LMIC RCTs mainly enrolled HCW with a mean age of around 40 years (Table 1). When limiting the analysis to the four RCTs which recruited HCWs (Europe and LMICs), the all-cause mortality RRR was 78% (−3-95%). In the five RCTs which recruited elderly and high-risk adults, the RRR was 43% (95% CI: −6-69%).

A different vaccine, a recombinant BCG variant (VPM1002), was used in one trial in Germany (44). Individuals vaccinated with VPM1002 had a higher mortality compared with placebo (6 vs 3 deaths; RR 2.00 (95% CI: 0.50 to 7.97)) than the trials comparing original BCG vaccines with placebo (18 vs 37 deaths; pooled RR 0.49 (95% CI: 0.28 to 0.85)) (test of same effect, p=0.065). In a sensitivity analysis including the VPM1002 trial with the other BCG vaccine trials (44), the all-cause mortality RRR was 40% (95% CI: 0.4 to64%).

### COVID-19 deaths

In the LMICs, half of the 10 placebo deaths were due to COVID-19. There were less COVID-19 deaths in the BCG groups compared with the placebo groups (0/1 vs 5/10) (Fisher’s exact test, p=0.06). In the European trials, COVID-19 deaths amounted to 30% (13/44) of the registered causes of death; COVID-19 deaths were similar in the BCG and placebo groups (6 vs 7 deaths) (Table 1). For all nine RCTs of BCG against COVID-19, BCG reduced the risk of death from COVID-19 by 50% (95% CI: −33 to 81%).

## Discussion

### Summary of the findings

Across nine RCTs, conducted during the COVID-19 pandemic, BCG compared with placebo in adults was associated with a marked reduction in all-cause mortality during the 6-12 months of follow-up.

It is interesting to compare this protective effect of BCG with that of mRNA vaccines. The two major RCTs of Pfizer and Moderna mRNA vaccines included 74,193 adults among whom there were 61 deaths. COVID-19 was responsible for 11% (7/61) of all deaths (28,29) and cardiovascular causes for 50% (27/54) (27–30). The estimated protective effect of BCG against COVID-19 death was comparable to the effect of mRNA vaccination (BCG: RR 0.50 (0.19 to1.33) vs. mRNA: 0.40 (0.08 to 2.06); p=0.82). In contrast, the estimated protective effect of BCG on all-cause mortality was greater than that of mRNA vaccines (BCG: RR 0.49 (0.28 to 0.85) vs mRNA: 1.03 (0.63 to 1.71); test of same effect, p=0.05) (Table 2).

**Table 2.**
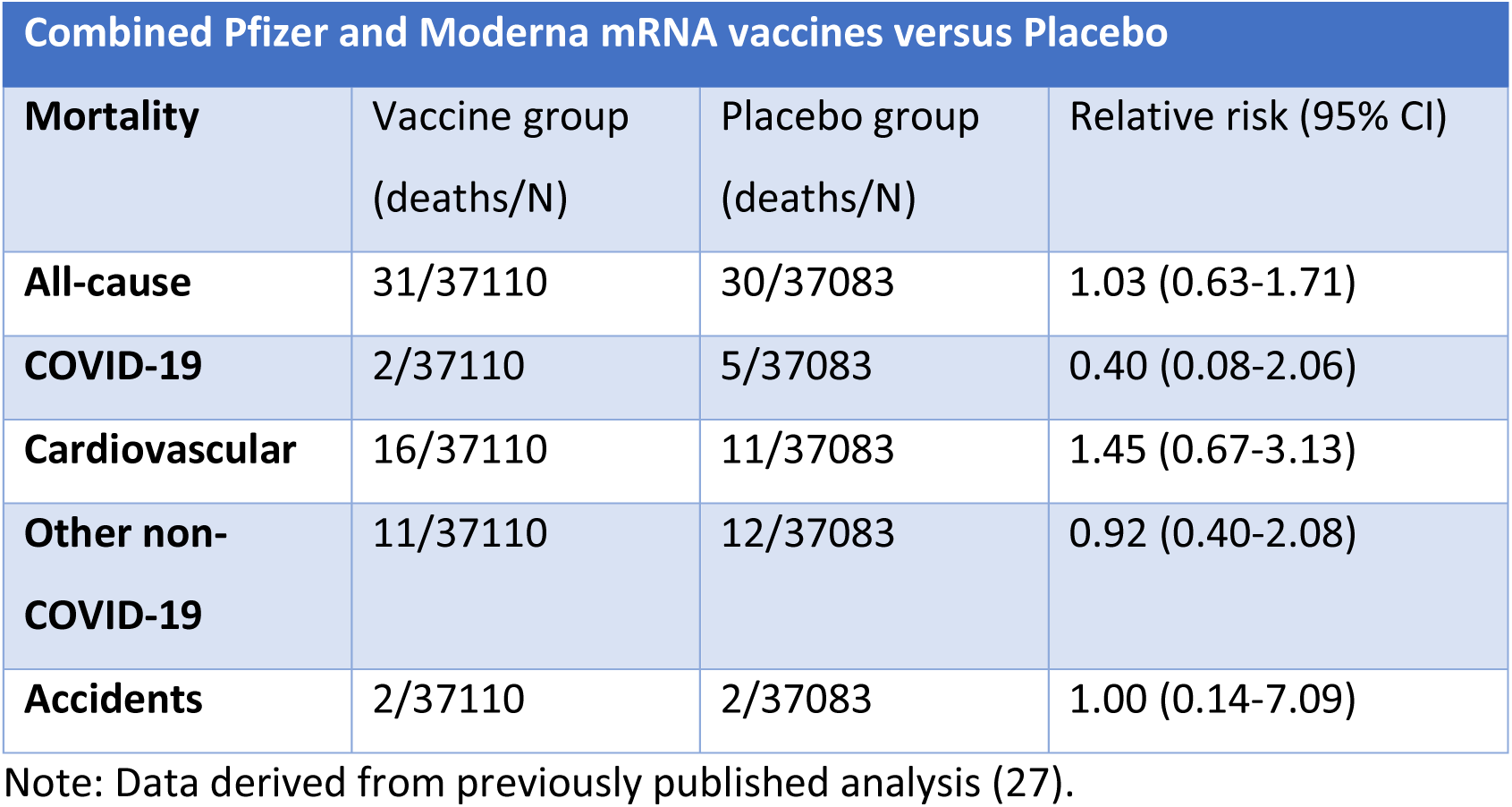
All-cause, COVID-19, and non-COVID-19 mortality in the RCTs of mRNA vaccines.

| **Combined Pfizer and Moderna mRNA vaccines versus Placebo** |  |  |  |
| --- | --- | --- | --- |
| **Mortality** | Vaccine group<br>(deaths/N) | Placebo group<br>(deaths/N) | Relative risk (95% CI) |
| **All-cause** | 31/37110 | 30/37083 | 1.03 (0.63-1.71) |
| **COVID-19** | 2/37110 | 5/37083 | 0.40 (0.08-2.06) |
| **Cardiovascular** | 16/37110 | 11/37083 | 1.45 (0.67-3.13) |
| **Other non-**<br>**COVID-19** | 11/37110 | 12/37083 | 0.92 (0.40-2.08) |
| **Accidents** | 2/37110 | 2/37083 | 1.00 (0.14-7.09) |

Note: Data derived from previously published analysis (27).

However, there could be important period effects or geographical differences that confound these comparisons. For example, the mortality level appeared to be higher in the placebo groups in the BCG trials (Table 1) than in the placebo groups in the mRNA trials (Table 2), most likely due to the higher age of the volunteers in most of the BCG trials, and our crude comparison could not be adjusted for trial differences, such as length of follow-up and age differences between participants in the RCTs.

Most of the BCG RCTs were done during a period when many individuals received specific COVID-19 vaccines during follow-up. Based on what is known from studies of non-specific effects in children (45,46) and supported by studies which investigated this, non-live vaccines given during follow-up may have modified the results (16,47,48). Unfortunately, information to study this aspect was not available.

### Consistency with other studies

To date, systematic reviews (SRs) in relation to BCG and COVID-19 have focused on the effect of BCG on COVID-19 incidence, hospitalisations, and ICU admission. They have not addressed the broader effect on all-cause outcomes. SRs and meta-analyses which include data on mortality are summarised in Supplementary Table 2; four of the five focus solely on COVID-19-related mortality, and one presented incomplete information on all-cause mortality (3 BCG vs 9 non-BCG deaths). While the previous SRs indicate effects in the same direction as the present meta-analysis, our data include many more events and details.

### Interpretation

Overall, the effect of BCG in terms of preventing COVID-19 was variable, ranging from very positive in some studies (12,30,40) to no or negative effects in other studies (13–16,18,19,24–26,37,38). Two RCTs among elderly with chronic underlying conditions in The Netherlands suggested that the BCG groups had more hospitalisations for cardiac arrhythmias (RR 2.0 (1.1-3.6)) (25). However, in the same two RCTs the BCG vs placebo RR for mortality was 0.77 (0.45-1.32). The two largest RCTs from LMICs (South Africa, Brazil) both had more severe COVID-19 episodes and hospitalisations in the BCG groups (18,19). However, in the same two RCTs there was lower mortality in the BCG groups, the combined estimate being 0 BCG deaths vs 8 placebo deaths (Fisher’s exact test, p=0.004). Thus, any negative effects of BCG on other outcomes does not seem to translate into negative effects on all-cause mortality.

Several potential explanations for the variation in BCG’s effects against COVID-19 have been proposed, including participant age, sex, comorbidity and vaccine strain (24,26), that too few BCG doses were given (in the studies that found a beneficial effect, participants had been primed with BCG before (12,16,40)), or that follow-up was too short (40). If BCG strengthens the innate immune response, (16,37) this could result in more episodes of disease and more hospitalisations being reported due to more severe symptoms induced by the immune responses (26). In both child and adult studies, this responsiveness may be amplified by subsequent vaccinations with non-live vaccines (37,40).

We have previously proposed that BCG might have a beneficial effect on adult mortality during the COVID-19 pandemic (2,11). To the four RCTs of BCG against COVID-19 included in a previous review (4), this meta-analysis adds five more RCTs with mortality data (15–17,19,20). The mortality reduction estimates were 0.54 (0.29 to 1.00) in the first four RCTs and 0.33 (0.09 to 1.23), in the five additional trials. To our knowledge there are no further RCTs of BCG against COVID-19 to be reported. Hence, the combined relative mortality risk of 0.49 (0.28 to 0.85) (Table 1) is the best estimate of how BCG might help to control a pandemic.

### Potential immunological mechanisms

Immunologically, the induction of protective heterologous effects against all-cause mortality by BCG vaccination could be induced through several potential mechanisms. First, BCG vaccination induces trained innate immunity in adults (14,51–54), resulting in modulation of antimicrobial innate host defence, including to SARS-CoV-2 (14,54). Second, BCG vaccination can also amplify the function of T-cells in an antigen-agnostic manner, a process also called heterologous T-cell immunity (55,56). Importantly, interaction between innate and adaptive immune responses are likely crucial for optimal antimicrobial activity, as shown by the amplification of trained immunity by lymphocyte-dependent interferon-gamma (57,58). Third, while BCG vaccination improves antimicrobial responsiveness, it can also dampen systemic inflammation in adults (54,59), which can result in improved adaptation to pathogens and better outcome of the infection, and also reduce the risk and severity of non-communicable diseases. A BCG-induced increase in innate host defence when infected, but reduced inflammation in the steady state, provides a potential immunological explanation for BCG reducing not only infectious mortality, but also mortality from non-communicable diseases (NCDs) during the pandemic, despite not specifically decreasing the risk of COVID-19 infection.

### Strength and limitations

The RCTs were not designed to investigate the overall effect on mortality. This is unlikely to affect the obtained estimates, but significant missing contextual information may have had an impact on the overall estimates. NSEs have often been observed to be sex-differential (1), but we were unable to test any effect of sex. The majority of HCWs were females and if the effect differs by sex, then this could have led to an overestimate or underestimate of the all-cause mortality effect.

Data on cause of death were missing for many deaths, potentially impacting results. However, any deaths from, for example, accidents and suicides are expected to be evenly distributed, mitigating bias.

Another limitation is that we used a fixed-effects meta-analysis, which assumes that the relative risk is independent of the setting. In the absence of a good understanding of the underlying mechanism by which BCG decreases all-cause mortality, a constant relative risk is hard to justify, and we cannot determine in which settings this effect applies. Subgroup analysis suggested that the effect is most pronounced in LMICs, although with more uncertainty because of the limited number of events in this setting.

### Future perspectives

The beneficial NSEs of BCG against mortality observed during a relatively short follow-up period warrant further studies. As the reduction in all-cause mortality was not limited to COVID-19 related deaths, such effects might have important public health consequences outside a pandemic. Therefore, RCTs should be done to confirm the putative reduction of all-cause mortality independent of a pandemic. Such trials could for example test the effect of BCG on seasonal influenza in adults in multi-country trials in both high and low-income settings (noteworthy, BCG may work better against influenza infection than against SARS-CoV2 infection (60)), and the effect of BCG on all-cause mortality in individuals of older age and in HCW in high-mortality settings.

From a global equity perspective, it particularly notable that although the protective effect of BCG on HCW mortality did not reach statistical significance due to small numbers, the point estimate suggests substantial benefit. In LMICs, where physician density is extremely low (e.g., ∼0.2/1000 in much of sub-Saharan Africa vs. 2.5-4 in Europe), each HCW is disproportionately valuable for health system resilience. Even a modest reduction in HCW deaths could translate into major system-level benefits, preventing the collapse of already fragile health systems, maintaining continuity of essential care, and reducing knock-on effects of HCW loss, such as increased neonatal mortality and uncontrollable outbreaks. Protecting HCWs in LMICs with BCG during pandemics should therefore be considered a high-priority strategy in pandemic preparedness and could also be considered a strategy outside pandemics.

In a global perspective, vaccine equity failed miserably during the COVID-19 pandemic. In a world mainly looking for disease specific and high-tech new vaccines, BCG could change the equity situation. If the beneficial effects of BCG are further supported, agencies for pandemic prevention should assure that the world has sufficient capacity to swiftly produce BCG vaccines when needed, and include deployment of BCG in pandemic preparedness plans. BCG is a cheap and safe vaccine and with the focus on vaccine equity in the Pandemic Treaty it is worth noting that it is already readily available in LMICs as a part of the Expanded Programme of Immunization.

## Conclusion

The current evidence suggests that BCG-vaccinated participants had reduced all-cause mortality in the trials done during the COVID-19 pandemic. BCG might be considered a potential interim approach to protect healthcare workers and the elderly during future pandemics, such as those caused by ‘Disease X’. The potential effects of BCG on all-cause mortality may also warrant further use of BCG in adult populations independent of a pandemic.

## Supporting information

PRISMA Checklist

## Contributions

PA, MGN and CSB developed the idea (4). All co-authors had taken part in or planned one or more of the original RCTs. SN and CHvW contributed to the statistical analysis. PA and FSB wrote the first draft of the manuscript, and all authors critically reviewed the manuscript, numerous times.

## Conflict of interest

CHvW reports research funding from DaVolterra, bioMeriéux, LimmaTech and Merck, unrelated to the current paper. The other authors have nothing to declare.

## Funding

There was no specific funding for the present analysis. The different RCTs had funding as reported in the original papers. The work of the Bandim Health Project on non-specific effects of vaccines has been supported by the University of Southern Denmark and Dr. Allan Schapira. NC is supported by a National Health and Medical Research Council (NHMRC) Investigator Grant (GNT1197117). LFP is supported by the Swiss National Science Foundation (Early Postdoc Mobility Grant number P2GEP3_178155 and Ambizione Grant number PZ00P3-209050).

## Independence

The funding agencies had no role in the study design, data collection, data analysis, data interpretation, or the writing of the report.

## Transparency declaration

The lead author affirms that the manuscript is an honest, accurate, and transparent account of the study being reported; that no important aspects of the study have been omitted.

## Data Availability

Data sharing: This meta-analysis used only data extracted from previously published studies. No new or individual participant data were generated. All underlying source data are available in the original trial reports and their supplementary materials. The complete dataset compiled for this meta-analysis, including all extracted variables used in the analyses, is submitted with the manuscript and will be deposited in a public repository and made openly available prior to publication. Code sharing: The full Stata code used for the meta-analysis is submitted as a supplementary file and will be deposited in a public repository and made openly available prior to publication.

https://zenodo.org/records/17649639

---

**Supplementary Table 1.**
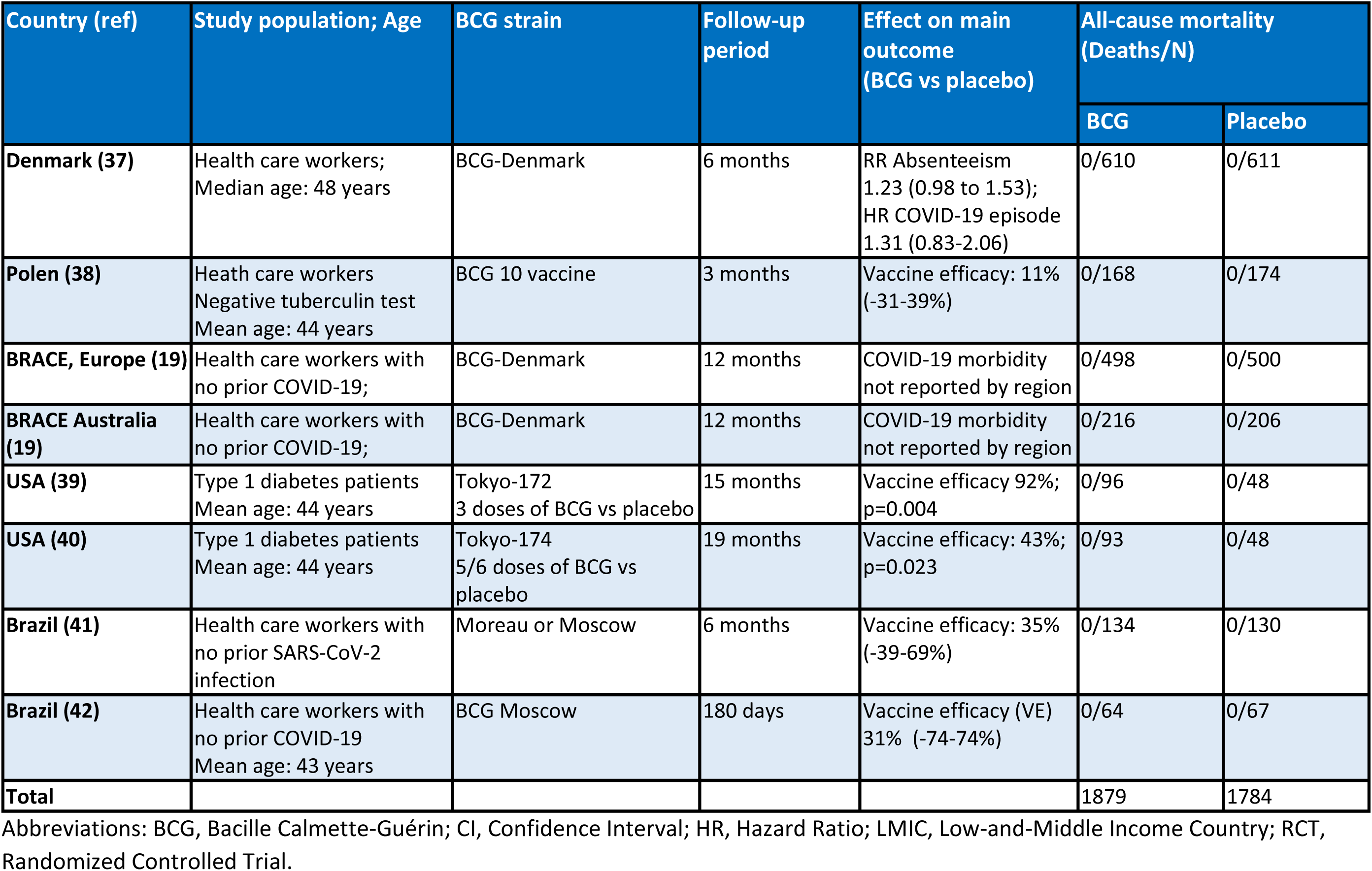
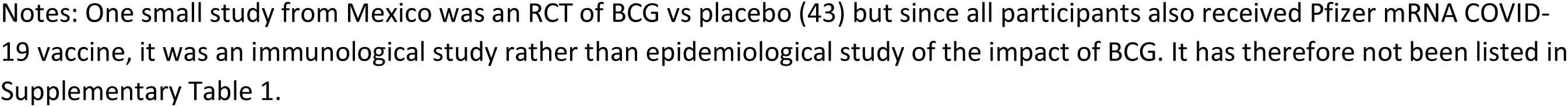
RCTs of BCG Vaccine versus Placebo against COVID-19 with no deaths during follow-up.

**Supplementary Table 2.**
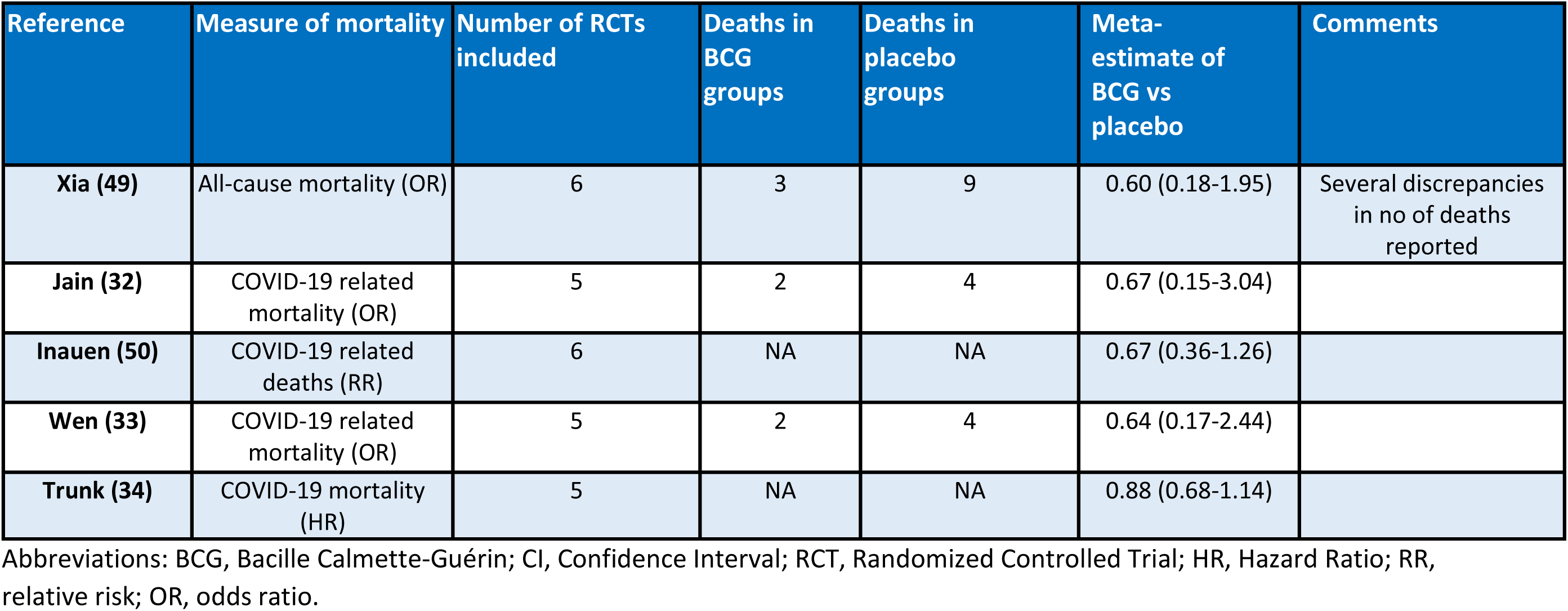
Meta-analyses of RCTs of BCG Vaccine versus Placebo against COVID-19.

**Supplementary figure 1.**
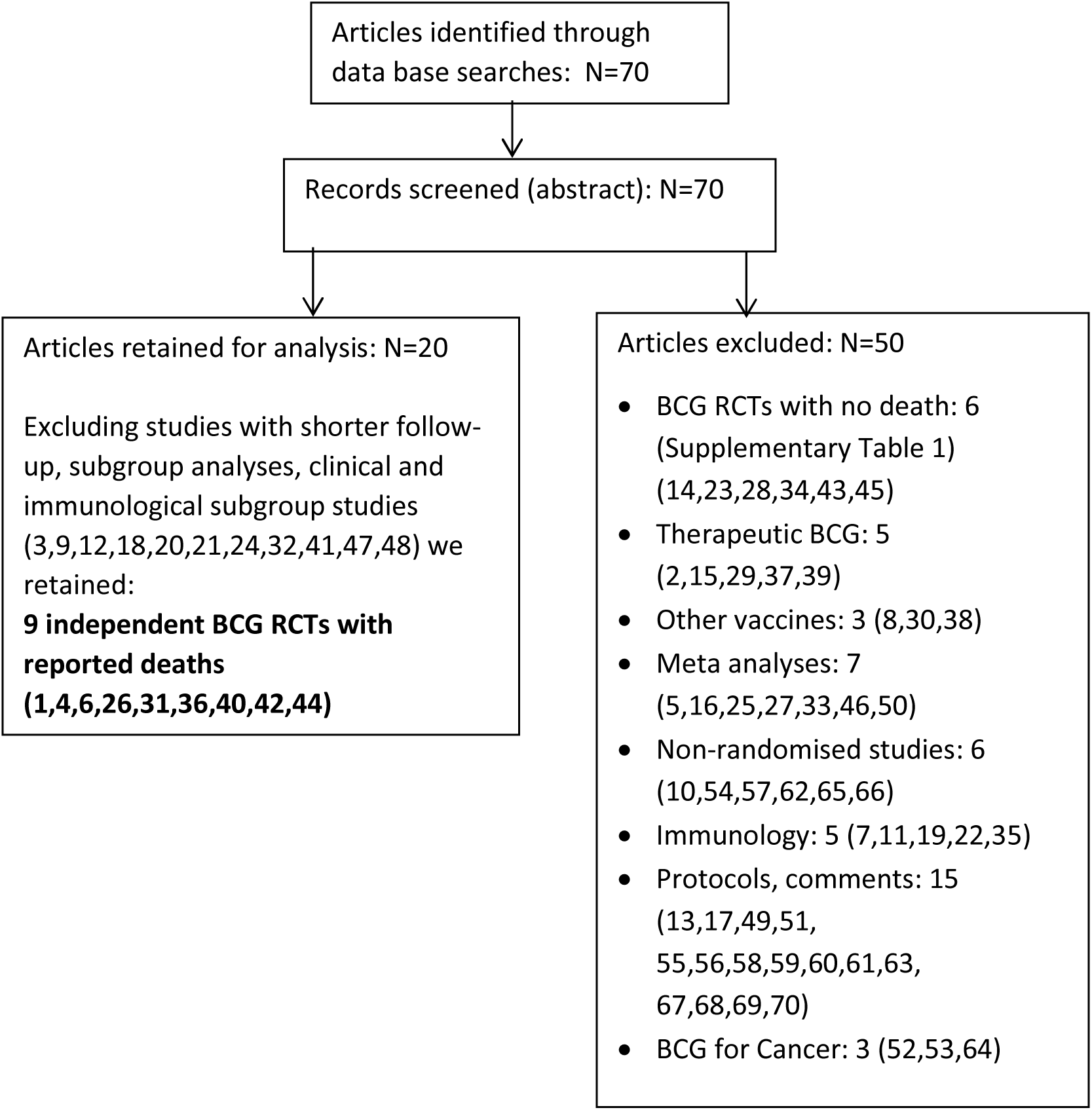
Systematic literature search for randomised trials comparing the effect BCG vs placebo on adult mortality

## Notes

### Competing Interest Statement

The authors have declared no competing interest.

### Funding Statement

This study did not receive any funding

### Author Declarations

This meta-analysis used only data extracted from previously published studies. No new or individual participant data were generated. All underlying source data are available in the original trial reports and their supplementary materials. All data received was de-identified.

