## Supplemental material for "The effect of BCG vaccination on adult mortality during the COVID-19 pandemic: a meta-analysis of randomised controlled trials"

**Supplementary Table 1. RCTs of BCG Vaccine versus Placebo against COVID-19 with no deaths during follow-up**

| Country (ref) | Study population; Age | BCG strain | Follow-up period | Effect on main outcome  (BCG vs placebo) | All-cause mortality (Deaths/N) | |
| --- | --- | --- | --- | --- | --- | --- |
|  |  |  |  |  | **BCG** | **Placebo** |
| Denmark (37) | Health care workers; Median age: 48 years | BCG-Denmark | 6 months | RR Absenteeism  1.23 (0.98 to 1.53);  HR COVID-19 episode 1.31 (0.83-2.06) | 0/610 | 0/611 |
| Poland (38) | Heath care workers  Negative tuberculin test  Mean age: 44 years | BCG 10 vaccine | 3 months | Vaccine efficacy: 11% (-31-39%) | 0/168 | 0/174 |
| BRACE, Europe (19) | Health care workers with no prior COVID-19; | BCG-Denmark | 12 months | COVID-19 morbidity not reported by region | 0/498 | 0/500 |
| BRACE, Australia (19) | Health care workers with no prior COVID-19; | BCG-Denmark | 12 months | COVID-19 morbidity not reported by region | 0/216 | 0/206 |
| USA (39) | Type 1 diabetes patients  Mean age: 44 years | Tokyo-172  3 doses of BCG vs placebo | 15 months | Vaccine efficacy 92%; p=0.004 | 0/96 | 0/48 |
| USA (40) | Type 1 diabetes patients  Mean age: 44 years | Tokyo-174  5/6 doses of BCG vs placebo | 19 months | Vaccine efficacy: 43%; p=0.023 | 0/93 | 0/48 |
| Brazil (41) | Health care workers with no prior SARS-CoV-2 infection | Moreau or Moscow | 6 months | Vaccine efficacy: 35% (-39-69%) | 0/134 | 0/130 |
| Brazil (42) | Health care workers with no prior COVID-19  Mean age: 43 years | BCG Moscow | 180 days | Vaccine efficacy (VE) 31% (-74-74%) | 0/64 | 0/67 |
| Total |  |  |  |  | 1879 | 1784 |

Abbreviations: BCG, Bacille Calmette-Guérin; CI, Confidence Interval; HR, Hazard Ratio; LMIC, Low-and-Middle Income Country; RCT, Randomized Controlled Trial.

Notes: One small study from Mexico was an RCT of BCG vs placebo (43) but since all participants also received Pfizer mRNA COVID-19 vaccine, it was an immunological study rather than epidemiological study of the impact of BCG. It has therefore not been listed in Supplementary Table 1.

**Supplementary Table 2. Meta-analyses of RCTs of BCG Vaccine versus Placebo against COVID-19**

| Reference | Measure of mortality | Number of RCTs included | Deaths in BCG groups | Deaths in placebo groups | Meta-estimate of BCG vs placebo | Comments |
| --- | --- | --- | --- | --- | --- | --- |
| Xia (49) | All-cause mortality (OR) | 6 | 3 | 9 | 0.60 (0.18-1.95) | Several discrepancies in no of deaths reported |
| Jain (32) | COVID-19 related mortality (OR) | 5 | 2 | 4 | 0.67 (0.15-3.04) |  |
| Inauen (50) | COVID-19 related deaths (RR) | 6 | NA | NA | 0.67 (0.36-1.26) |  |
| Wen (33) | COVID-19 related mortality (OR) | 5 | 2 | 4 | 0.64 (0.17-2.44) |  |
| Trunk (34) | COVID-19 mortality (HR) | 5 | NA | NA | 0.88 (0.68-1.14) |  |

Abbreviations: BCG, Bacille Calmette-Guérin; CI, Confidence Interval; RCT, Randomized Controlled Trial; HR, Hazard Ratio; RR, relative risk; OR, odds ratio.

**Supplementary Figure 1. Systematic literature search for randomised trials comparing the effect BCG vs placebo on adult mortality**

Articles identified through data base searches: N=70

Records screened (abstract): N=70

Articles excluded: N=50

- BCG RCTs with no death: 6 (Supplementary Table 1) (14,23,28,34,43,45)
- Therapeutic BCG: 5 (2,15,29,37,39)
- Other vaccines: 3 (8,30,38)
- Meta analyses: 7 (5,16,25,27,33,46,50)
- Non-randomised studies: 6 (10,54,57,62,65,66)
- Immunology: 5 (7,11,19,22,35)
- Protocols, comments: 15 (13,17,49,51, 55,56,58,59,60,61,63, 67,68,69,70)
- BCG for Cancer: 3 (52,53,64)

Articles retained for analysis: N=20

Excluding studies with shorter follow-up, subgroup analyses, clinical and immunological subgroup studies (3,9,12,18,20,21,24,32,41,47,48) we retained:

**9 independent BCG RCTs with reported deaths (1,4,6,26,31,36,40,42,44)**

Notes: Search: ((randomized or randomised) trial or RCT) and (BCG or Bacille Calmette Guerin) and (COVID-19 or SARS-CoV-2), restricted to the years 2020-2024 [December 2024]
